# Hydraulic forces assist left ventricular filling in aortic stenosis and improve after valve replacement in patients with the lowest hydraulic force and otherwise healthiest left ventricles at baseline

**DOI:** 10.1101/2025.06.19.25329899

**Authors:** Bryce Watson, Jonathan Bennett, Nikoo Aziminia, Abhishek Shetye, George D Thornton, Rhodri Davies, Iain Pierce, Rebecca Kozor, Alun D Hughes, James C Moon, Martin Ugander, Thomas A Treibel

## Abstract

**Background:** Diastolic dysfunction in the setting of aortic valve replacement (AVR) for aortic stenosis (AS) is incompletely understood. This study aims to to assess the net hydraulic force of left ventricular (LV) filling in participants with severe symptomatic AS undergoing AVR.

**Methods:** This single-centre prospective observational cohort study evaluated patients with severe, symptomatic AS undergoing AVR between 2012-2015. Clinical assessment and cardiovascular magnetic resonance (CMR) was completed prior to AVR and 1-year post-operatively. Atrioventricular area difference (AVAD) was used as a surrogate for the hydraulic force of LV filling. AVAD at mid-diastole was measured as the difference between LV short-axis area and left atrial short-axis area.

**Results:** In patients with AS (n=110, 54% male, age 71 [64–77] years, aortic valve area 0.74±0.25 cm^2^), AVAD was positive at baseline (2.8±6.5 cm^2^) consistent with a net hydraulic force assisting LV filling. While AVAD did not change post-operatively on a group level (p=0.70), an improvement in AVAD was associated univariably with increasing baseline LV ejection fraction, and decreasing baseline AVAD, LV volume, mass, myocardial extracellular volume, and infarct size (p<0.05 for all), and multivariably with baseline decreasing AVAD, LV mass, and age (model adjusted R^2^=0.49, p<0.001).

**Conclusion:** In severe AS, hydraulic force contributes to LV filling prior to and following AVR. The greatest improvement in hydraulic force following AVR occurred in those with the lowest baseline hydraulic force, but also with lower age and the absence of otherwise deleterious LV myocardial remodelling, thus supporting the benefits of early intervention.

## INTRODUCTION

Aortic Stenosis (AS) is the most common valvular heart disease (VHD) in developed countries and is becoming increasingly more prevalent due to aging populations[1]. Given the prevalence in elderly individuals, definitive treatment of AS with either open surgical or transcatheter aortic valve replacement (AVR)[2] merits careful consideration. Half of all individuals with AS, and two thirds of those having AVR for severe AS have diastolic dysfunction[3,4]. Impaired diastolic function pre-operatively has been demonstrated to be a predictor of poor outcomes including mortality following AVR[3,4,5]. A meta-analysis has shown higher all-cause mortality and major adverse cardiovascular events following transcatheter AVR in those with severe diastolic dysfunction[6]. This relationship appears to be dependent on severity with those stratified into higher grades of diastolic dysfunction pre-operatively having increased rates of cardiovascular death or hospitalization following AVR[7].

Reverse remodelling has been demonstrated following AVR with reduction in left ventricular (LV) mass and regression of diffuse myocardial fibrosis seen in the first year following intervention[8,9,10,11]. However, this reverse remodelling does not necessarily lead to improvement in diastolic function[12]. The echocardiographic grade of diastolic dysfunction pre-operatively has been shown to improve following AVR in between 50-71% of cases[13,14]. Those whose grade of diastolic dysfunction improves following AVR have lower rates of death and hospitalisation at 1 and 2 years post-intervention[13]. This lack of improvement in diastolic function in some individuals despite improvements in myocardial hypertrophy and diffuse myocardial fibrosis is incompletely understood and warrants further investigation.

The forces that contribute to diastolic LV filling include active relaxation[15], elastic restorative forces[16], left atrial contraction[17], and hydraulic force[18]. The hydraulic force of LV filling is determined by the geometric relationship between the LA and LV[18]. Of note, the hydraulic force of LV filling is yet to be assessed in individuals with AS or in those having AVR. It is not known whether the remodelling and reverse remodelling seen in AS will impact the hydraulic force. Hydraulic forces have been shown to provide independent prognostic information beyond diastolic dysfunction grading[19], and may provide insights into diastolic dysfunction in AS, particularly persisting diastolic dysfunction following AVR. This study aims to analyze the atrioventricular area difference (AVAD) as a surrogate for the hydraulic force of LV filling in participants with severe AS before and after AVR and investigates its relationship with known markers of myocardial function and tissue remodelling.

## METHODS

This is a sub-study of the prospective RELIEF-AS study of patients who had AVR for severe AS between January 2012 and January 2015 (NCT02174471)[8]. The participants were from a single referral centre (University College London Hospitals NHS Trust, London) and were recruited prior to having their intervention. This study was approved by the United Kingdom National Research Ethics Service (19/LO/1849). The participants completed clinical, biochemical, 6-minute walk test (6MWT)[20], echocardiographic, and cardiovascular magnetic resonance (CMR) assessments at baseline and 1-year following AVR.

### Study sample

Participants were individuals with severe symptomatic AS who underwent AVR with or without coronary artery bypass graft (CABG). Those over the age of 18 with severe AS were included. Severe AS was defined as two or more of: aortic valve area (AVA) <1cm^2^, velocity time integral ratio <0.25, aortic valve peak pressure gradient ≥64mmHg, mean pressure gradient ≥40mmHg, or confirmation of severe AS using an alternative imaging modality. Those with severely impaired renal function (estimated glomerular filtration rate <30ml/min/1.73m^2^), previous cardiac valve procedures, infective endocarditis, severe mitral regurgitation (MR), severe aortic regurgitation (AR), cardiac amyloid, insufficient CMR acquisitions for AVAD analysis, or those pregnant or breast feeding were excluded from the study.

### Cardiovascular Magnetic Resonance

CMR was performed using a standard clinical protocol at 1.5 Tesla (Magnetom Avanto, Siemens Healthineers, Germany). Late gadolinium enhancement (LGE), and T1 mapping by Modified Look-Locker Inversion recovery were performed before and after administration of gadolinium contrast (gadolinium-DOTA, Guerbet S.A., France). LGE imaging was completed 10 minutes post contrast administration and analysed as regions with a signal >3SD from remote. T1 mapping was performed before and 15 minutes post contrast for calculating myocardial extracellular volume fraction (ECV). Images were analysed on CVI42 software version 5.12 (Circle Cardiovascular Imaging, Canada). Global thickness (GT) was calculated using the equation: GT = 0.05 + (1.60 x LVM[g]^0.84^ x LVEDV[ml]^−0.49^) [21].

### Atrioventricular area difference analysis

Endocardial ventricular short-axis area (VSA) measurement was performed with the assistance of a validated machine learning model[22]. The model identified the endocardial contours to measure the VSA and was manually corrected as needed. Papillary muscles were included in the blood volume, and CMR slices including the left ventricular outflow tract were excluded.

Atrial short-axis area (ASA) was assessed using a circular approximation method from the mean of 3 manually measured maximum LA diameters. The short-axis acquisitions could not be used to measure the ASA as they incompletely imaged the base of the LA and therefore may not have included the maximum ASA. Diameter measurements were obtained on 4-chamber, 3-chamber and 2-chamber long axis acquisitions. The maximum short-axis LA diameters on each acquisition were measured perpendicular to the long axis of the left ventricle (Figure 1). All measurements were made at the same distance from the mitral valve annulus within 5mm of each other. The ASA was calculated from the mean of the maximum atrial short-axis diameters using circular approximation calculated from the radius of the short-axis measurements.

**Figure 1:**
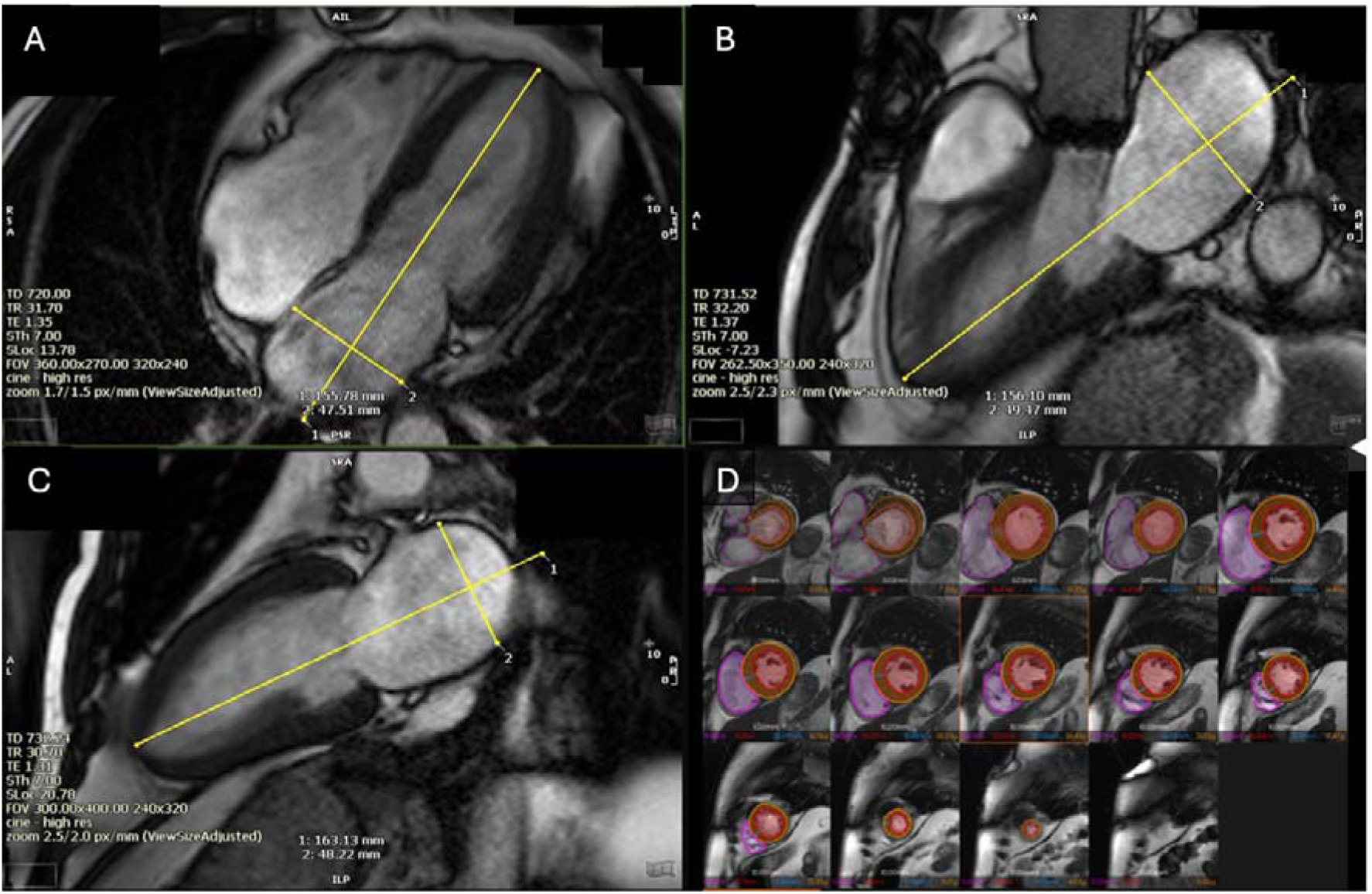
Atrioventricular Area Difference (AVAD) methodology. Atrial short-axis area (ASA) was measured using mean maximum left atrial diameters on long axis acquisitions and circular approximation calculation. Maximum left atrial diameters were measured perpendicular to long axis of left ventricle (A,B,C). Ventricular short-axis area (VSA) was measured from short-axis acquisitions with a validated machine learning model (D). Atrioventricular area difference (AVAD) was calculated as VSA minus ASA.

AVAD was used as a surrogate measurement for the net hydraulic force acting on the AV plane, and was measured on CMR at mid-diastole as follows:

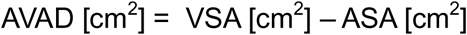

Change in AVAD (ΔAVAD) was calculated as post-operative AVAD minus pre-operative AVAD.

### Echocardiography

Transthoracic echocardiography was performed using Vivid E9 system (GE Healthcare, Wisconsin, USA) and measurements were performed according to the guidelines of the American Society of Echocardiography and European Society of Echocardiography [23].

### Biochemistry

Enzyme Linked Immunosorbent Assay was used to analyse serum N-terminal pro-brain natriuretic peptide (NT-proBNP) and high sensitivity troponin T (hsTnT) (Roche Diagnostics, Indiana, USA).

### Statistical Analysis

Statistical analysis was performed on STATA-MP Version 18.0 (StataCorp, Texas, USA). Normality was assessed with quantile-quantile plots and Shapiro-Wilk tests. Unpaired t-tests were used to compare means between two groups of normally distributed continuous variables and paired t-tests if these variables were paired. Pearson correlation coefficients were used to assess linear correlations between two continuous variables and Spearman non-parametric rank correlation coefficients for monotonic non-linear relationships between continuous variables. Multivariable linear regression was used to adjust correlations for potential confounders. Confounders were chosen based on prior evidence that they might be common causes of the exposure and outcome. Highly skewed variables were log transformed prior to inclusion in models. All analysis were based on complete case data. Missing data was addressed using listwise deletion.

## RESULTS

### Baseline study population

161 participants completed pre-operative assessment including CMR. 51 were excluded from the study prior to analysis; this was due to being managed medically, identification of concurrent cardiac amyloid, Fabry Disease, severe MR or pseudo-severe AS, as well as claustrophobia, being acutely unwell, cardiac pacemaker insertion, having inadequate CMR acquisitions available or death prior to post-operative CMR (Figure 2). The median age of the 110 included patients was 71 years and 54% were male. LVEF was 70 [64–80] %, peak aortic valve velocity 4.4±0.6m/s and mean aortic valve gradient 47 ±14 mmHg (Table 1).

**Figure 2:**
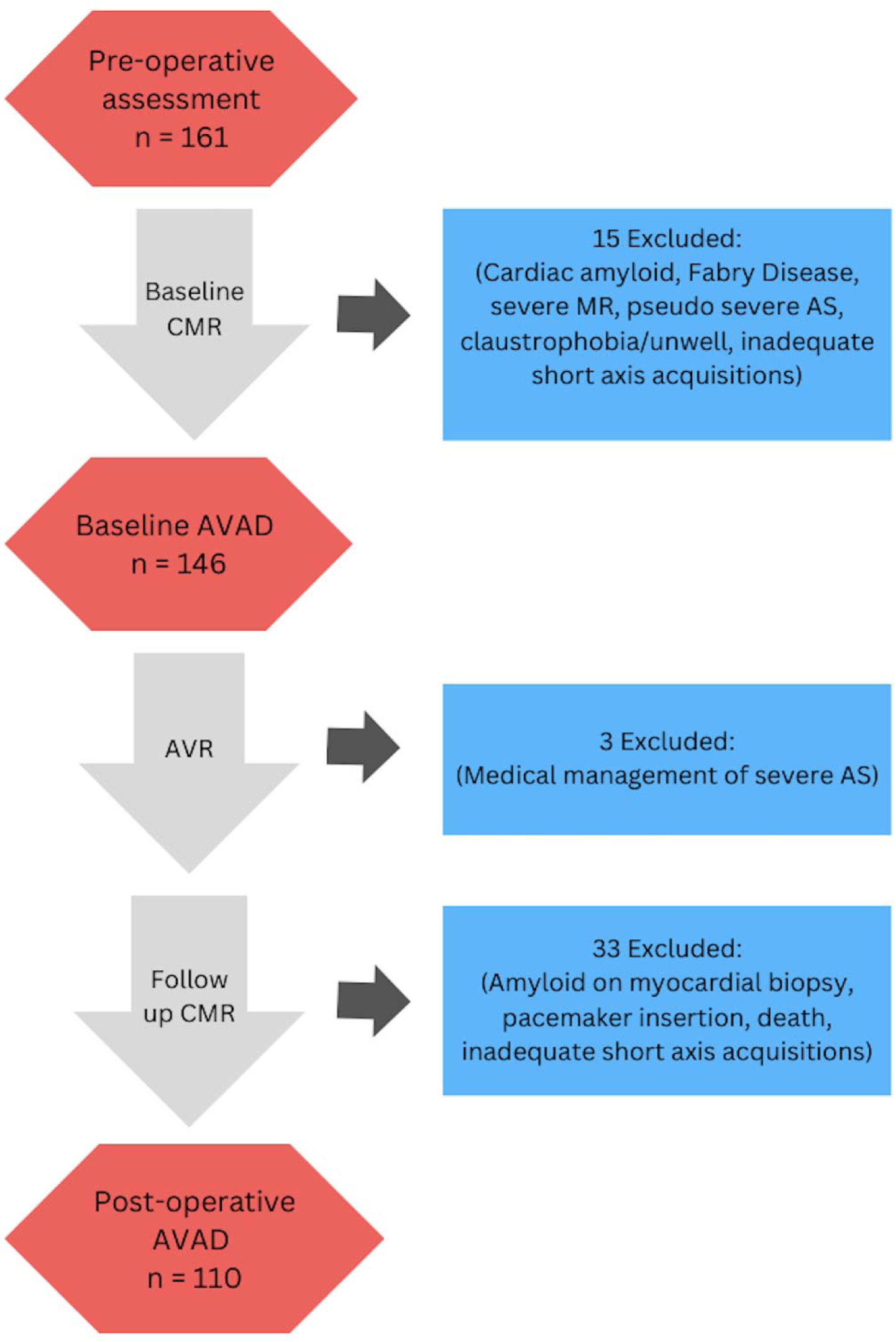
Study consort diagram CMR = cardiovascular magnetic resonance; AS = aortic stenosis; MR = mitral regurgitation; AVAD = atrioventricular area difference; AVR = aortic valve replacement

**Figure 3:**
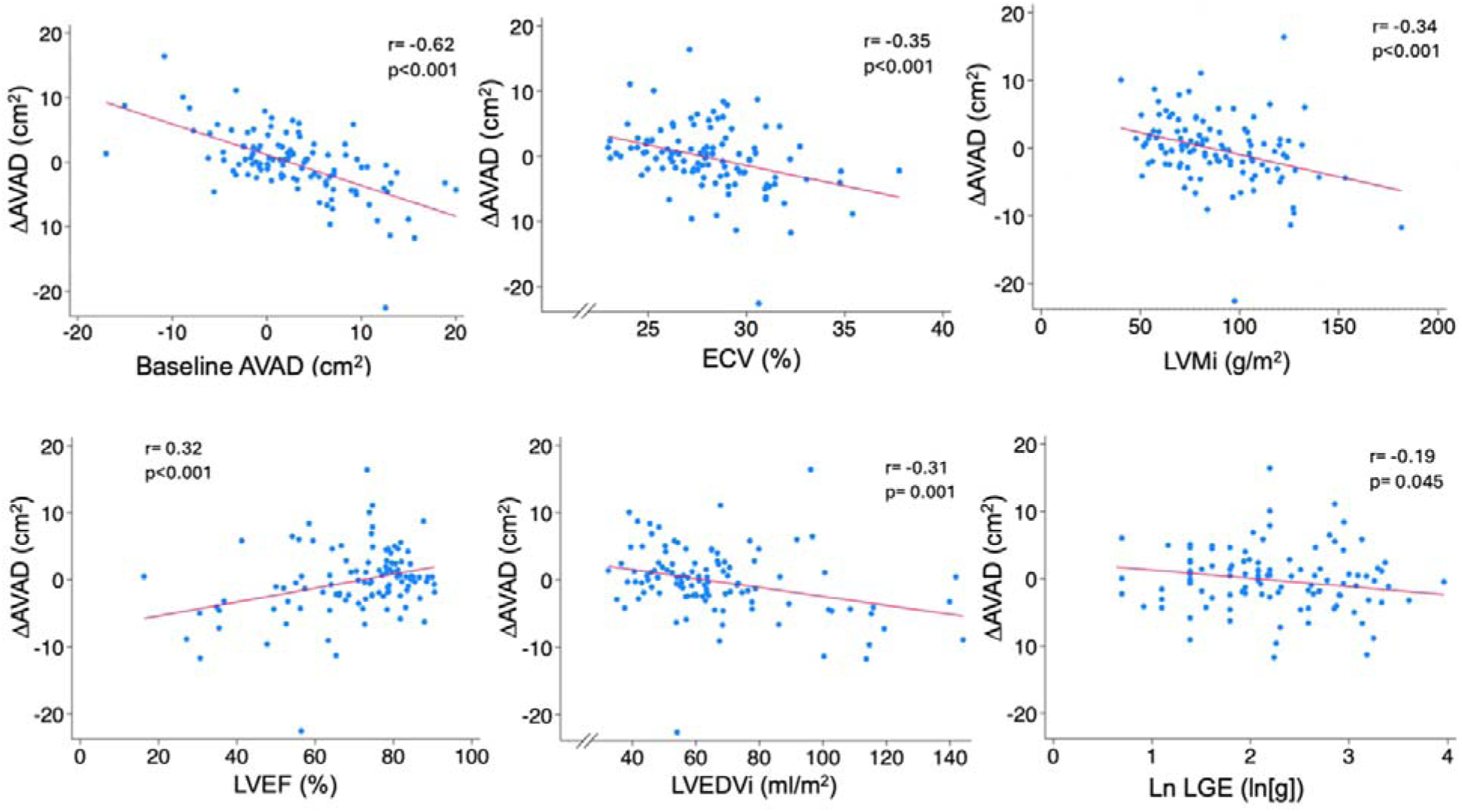
ΔAVAD correlations with baseline parameters. ΔAVAD = change in atrioventricular area difference (cm^2^); ECV = left ventricular extracellular volume (%); LVMi = Left Ventricular Mass index (g/m^2^); LVEF = left ventricular ejection fraction (%); LVEDVi = left ventricular end diastolic volume (ml/m^2^); LGE = late gadolinium enhancement (g).

**Table 1:**
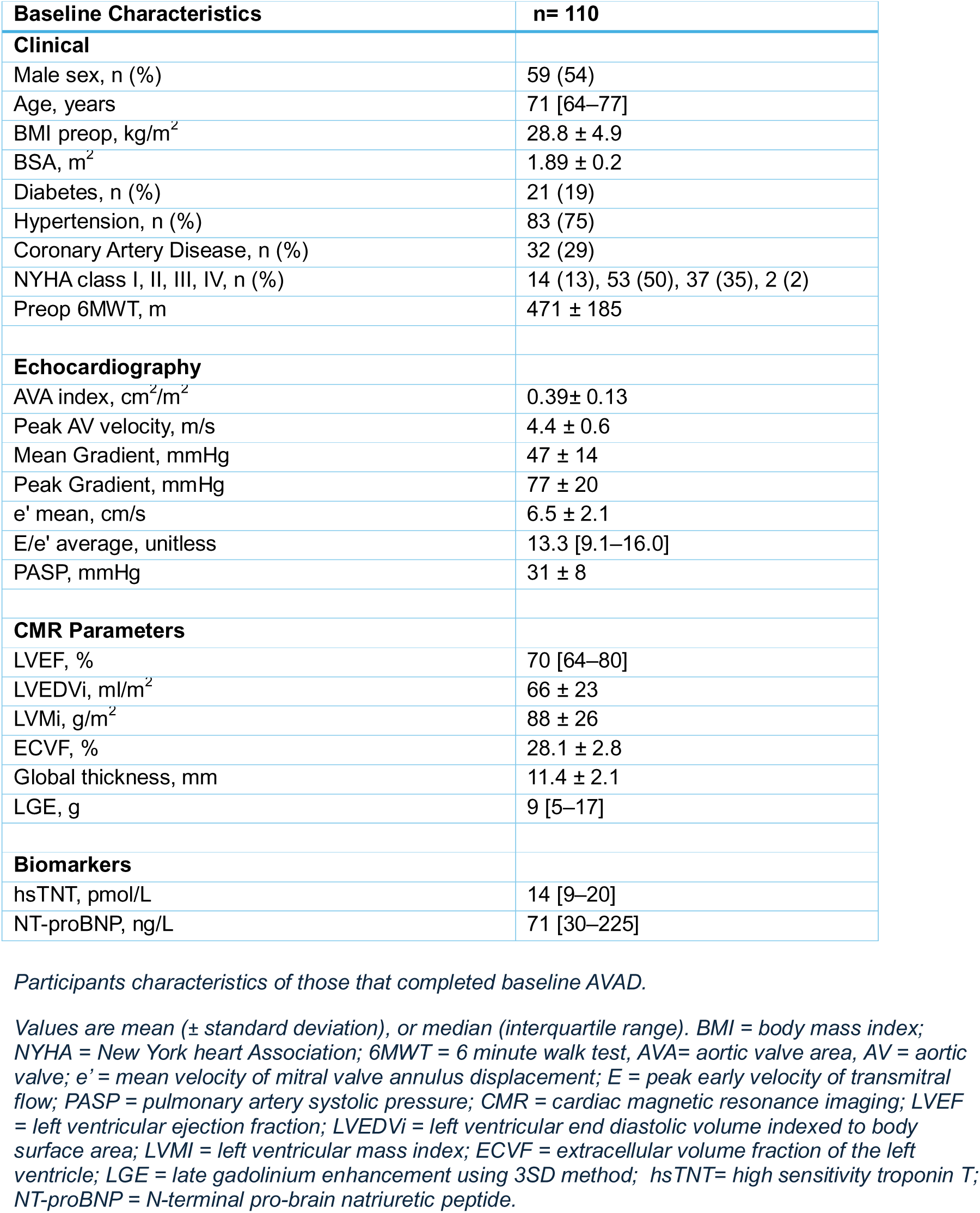
Baseline participant characteristics.

| Baseline Characteristics | n= 110 |
| --- | --- |
| **Clinical** |  |
| Male sex, n (%) | 59 (54) |
| Age, years | 71 [64–77] |
| BMI preop, kg/m<sup>2</sup> | 28.8 ± 4.9 |
| BSA, m<sup>2</sup> | 1.89 ± 0.2 |
| Diabetes, n (%) | 21 (19) |
| Hypertension, n (%) | 83 (75) |
| Coronary Artery Disease, n (%) | 32 (29) |
| NYHA class I, II, III, IV, n (%) | 14 (13), 53 (50), 37 (35), 2 (2) |
| Preop 6MWT, m | 471 ± 185 |
| **Echocardiography** |  |
| AVA index, cm<sup>2</sup>/m<sup>2</sup> | 0.39± 0.13 |
| Peak AV velocity, m/s | 4.4 ± 0.6 |
| Mean Gradient, mmHg | 47 ± 14 |
| Peak Gradient, mmHg | 77 ± 20 |
| e' mean, cm/s | 6.5 ± 2.1 |
| E/e' average, unitless | 13.3 [9.1–16.0] |
| PASP, mmHg | 31 ± 8 |
| **CMR Parameters** |  |
| LVEF, % | 70 [64–80] |
| LVEDVi, ml/m<sup>2</sup> | 66 ± 23 |
| LVMi, g/m<sup>2</sup> | 88 ± 26 |
| ECVF, % | 28.1 ± 2.8 |
| Global thickness, mm | 11.4 ± 2.1 |
| LGE, g | 9 [5–17] |
| **Biomarkers** |  |
| hsTNT, pmol/L | 14 [9–20] |
| NT-proBNP, ng/L | 71 [30–225] |

*Participants characteristics of those that completed baseline AVAD.*
*Values are mean (± standard deviation), or median (interquartile range). BMI = body mass index; NYHA = New York heart Association; 6MWT = 6 minute walk test, AVA= aortic valve area, AV = aortic valve; e' = mean velocity of mitral valve annulus displacement; E = peak early velocity of transmitral flow; PASP = pulmonary artery systolic pressure; CMR = cardiac magnetic resonance imaging; LVEF = left ventricular ejection fraction; LVEDVi = left ventricular end diastolic volume indexed to body surface area; LVMi = left ventricular mass index; ECVF = extracellular volume fraction of the left ventricle; LGE = late gadolinium enhancement using 3SD method; hsTNT= high sensitivity troponin T; NT-proBNP = N-terminal pro-brain natriuretic peptide.*

### Baseline AVAD

Baseline AVAD was 2.8±6.5cm^2^ consistent with a net positive hydraulic force assisting LV filling. AVAD did not differ between sexes (males 3.8±7.0cm^2^ vs females 1.7±5.7cm^2^, p=0.09). Baseline AVAD was negatively correlated with age (rho=-0.40, p<0.001).

### Baseline AVAD correlations

Correlations between baseline AVAD and baseline clinical, biochemical, echocardiographic and CMR variables are summarized in Table 2. The strongest correlations were between baseline AVAD and baseline LV end-diastolic volume index (LVEDVi) (r=0.58, p<0.001), LVEF (r=-0.48, p<0.001), and LV mass index (LVMi) (r=0.46, p<0.001). Correlations were also present between baseline AVAD and age, diffuse myocardial fibrosis (ECV) and focal myocardial fibrosis (LGE).

**Table 2:** Baseline AVAD correlations with baseline variables.

| Baseline Variable | Correlation (r) | Comparison of means | p-value |
| --- | --- | --- | --- |
| Age | **-0.39** † | - | **<0.001** |
| Sex | - | Males: $3.8 \pm 7.0 \text{ cm}^2$<br>Females: $1.7 \pm 5.7 \text{ cm}^2$ | 0.09 |
| Coronary Artery Disease | - | Present $3.4 \pm 8.1 \text{ cm}^2$<br>Absent $2.5 \pm 5.8 \text{ cm}^2$ | 0.51 |
| Hypertension | - | Present $2.4 \pm 6.6 \text{ cm}^2$<br>Absent $4.0 \pm 6.1 \text{ cm}^2$ | 0.28 |
| Baseline 6MWT | 0.19 | - | 0.07 |
| ln(hsTNT) | **0.22** | - | **0.02** |
| ln(NT-proBNP) | **0.25** | - | **0.01** |
| e' | -0.06 | - | 0.58 |
| E/e' | **0.18** | - | **0.09** |
| LVEF | **-0.48** | - | **<0.001** |
| LVEDVi | **0.58** | - | **<0.001** |
| LVMi | **0.46** | - | **<0.001** |
| Ln LGE | **0.22** | - | **0.02** |
| ECV | **0.34** | - | **<0.001** |

*Correlations between baseline Atrioventricular Area Difference and baseline clinical, biochemical and* *imaging variables, n =110.*
*Pearson's correlation used for parametric data and Spearman's Rank correlation (†) for non-* *parametric data. 6MWT = 6 minute walk test, hsTNT= high sensitivity troponin T; NT-proBNP = N-* *terminal pro-brain natriuretic peptide; e' = mean velocity of mitral valve annulus displacement; E =* *peak early velocity of transmitral flow; LVEF = left ventricular ejection fraction; LVEDVi = left ventricular* *end diastolic volume indexed to body surface area; LVMi = left ventricular mass index; LGE 3SD = late* *gadolinium enhancement using 3SD method; ECV Fraction = extracellular volume fraction of the left* *ventricle.*

### Intervention

5 participants had TAVI (5%), 68 had tissue SAVR (62%), 33 had mechanical SAVR (30%) and 4 had suture-less SAVR (4%). 35 participants (32%) had concomitant coronary artery bypass grafting (CABG).

### Associations with changes in AVAD

Correlations between change in AVAD (ΔAVAD) and baseline clinical, biochemical, echocardiographic and CMR variables were assessed with key findings summarized in Table 3. In univariable analysis there were negative correlations between ΔAVAD and ECV, LVMi, LVEDVi, NT-pro-BNP, and hsTNT. Lower baseline values of these variables were associated with a larger increase in AVAD after AVR. The strongest correlation was a negative correlation between ΔAVAD and baseline AVAD. In a multivariable model adjusting for potential confounders, baseline AVAD (standardized beta = -0.65, p<0.001), age, and LVMi were independently associated with ΔAVAD. The overall multivariable model had an adjusted R^2^=0.49 (p<0.001). When comparing ΔAVAD between those whose baseline AVAD was below or above the median of baseline AVAD (2.4 cm^2^), those below the median had a greater ΔAVAD (2.1±4.0 vs -2.5±5.0cm^2^, p<0.001). Baseline and post-operative AVAD did not differ overall (2.8±6.5 vs 2.6±5.2cm^2^, p=0.70).

**Table 3:** Correlations between ΔAVAD and baseline variables.

| Baseline variable | Correlation coefficient | Difference in means | Univariable p-value | Standardized Multivariable coefficient | Multivariable p-value |
| --- | --- | --- | --- | --- | --- |
| Male sex |  | +0.67cm<sup>2</sup> | 0.485 | 0.07 | 0.416 |
| Age, years | 0.01 | - | 0.887 | **-0.26** | **0.003** |
| 6MWT, meters | 0.05 | - | 0.621 | - | - |
| ln(hsTNT), ln(ng/mL) | **-0.31** | - | **0.001** | - | - |
| ln(NT-proBNP), ln(pg/ml) | **-0.33** | - | **<0.001** | - | - |
| e', cm/s | 0.06 | - | 0.594 | - | - |
| E/e', unitless | -0.20 | - | 0.072 | - | - |
| LVEF, % | **0.32** | - | **<0.001** | 0.01 | 0.923 |
| LVEDVi, ml/m<sup>2</sup> | **-0.31** | - | **0.001** | 0.25 | 0.053 |
| LVMi, g/m<sup>2</sup> | **-0.34** | - | **<0.001** | **-0.28** | **0.021** |
| ln(LGE), ln(g) | **-0.19** | - | **0.045** | 0.02 | 0.057 |
| ECV, % LV | **-0.35** | - | **<0.001** | -0.15 | 0.071 |
| Baseline AVAD, cm<sup>2</sup> | **-0.62** |  | **<0.001** | **-0.65** | **<0.001** |
|  |  |  |  | **Multivariable model**<br>**adjusted R<sup>2</sup> = 0.49, p<0.001** |  |

*Correlations between change in AVAD ( ΔAVAD) and baseline clinical, biochemical and imaging* *variables.*
*Multivariable model included sex, age, LVEF, LVEDVi, LVMi, Ln(LGE), ECV.*
*6MWT = 6 minute walk test, hsTNT= high sensitivity troponin T; NT-proBNP = N-terminal pro-brain* *natriuretic peptide; LVEF = left ventricular ejection fraction; LVEDVi = left ventricular end diastolic* *volume indexed to body surface area; e' = mean velocity of mitral valve annulus displacement; E =* *peak early velocity of transmitral flow; LVMi = left ventricular mass index; LGE = late gadolinium* *enhancement using 3SD method; ECV Fraction= extracellular volume fraction of the left ventricular* *myocardium.*

### Echocardiographic diastolic function

Echocardiographic diastolic function improved following AVR. E/e’ decreased (12.5 [9.1-16.0] vs 10.3 [7.8-12.6], p<0.01) and e’ increased (6.6±2.1 vs 8.1±2.7 cm/s, p<0.01). No correlation was found between baseline AVAD or ΔAVAD and baseline E/e’ or e’ velocity (p>0.05 for all).

### Functional Outcome

No convincing correlation was found between baseline 6MWT and either baseline AVAD (r=0.19, p=0.07) or ΔAVAD (r=0.05, p=0.62). However, post-operative AVAD was correlated with post-operative 6MWT (r=0.39, p<0.01).

## DISCUSSION

This was the first study assessing the hydraulic force of LV filling in AS and following AVR. The study demonstrated that AVAD, as a surrogate for the hydraulic force, was positive at baseline and post-operatively, therefore assisting LV filling in both instances. While AVAD did not change overall following AVR, improvement in AVAD associated inversely with baseline AVAD, and this association persisted in multivariable analysis. Notably, improvement in AVAD associated with lower age and the absence of otherwise deleterious LV myocardial remodelling (increased LVMI & ECV), possibly supporting the benefits of early intervention.

A positive AVAD at baseline and following AVR is consistent with a net hydraulic force acting on the AV plane in an apex-to-base direction which assists LV filling in AS. This novel physiological process occurs in addition to active relaxation, elastic restorative forces, and left atrial contraction. Previous studies have also found a net positive hydraulic force in pathologies such as heart failure with reduced ejection fraction [24] and atrial septal defects (ASDs)[25].

On average in this study, the AVAD did not change following AVR. This contrasts with studies investigating AVAD in other pathologies and interventions. In a study assessing the hydraulic force before and after closure of atrial septal defect an increase in AVADi (AVAD indexed to body surface area) from a median of 6.4 cm/m2 to 8.7 cm/m^2^ was observed[25]. ASD closure changes the volume loading between the heart chambers and therefore impacts left atrial and left ventricular size and consequently AVAD. Changes in volume loading, such as due to atrial septal defects may cause more dynamic changes in chamber size than interventions for pressure loading conditions like AS [26,27]. This is a potential explanation for why the mean hydraulic force did not change discernibly in this study.

In contrast, pressure loading pathologies have been shown to cause a greater degree of LA enlargement than volume loading pathologies[27]. This knowledge inspired the hypothesis of the current study, namely that AVAD would increase post AVR, due to a decreased LA size after intervention. However, this study did not demonstrate a decrease in ASA or LA size following AVR. A 2024 meta-analysis of 1066 patients assessing LA size following TAVI demonstrated only a modest 2.7ml/m^2^ reduction in LA volume indexed to body surface area (LAVi) [28]. This is only a 5% reduction in volume from baseline and such a small change may not have been detectable in our study. In general, there is a paucity of research investigating how the hydraulic force changes with different valvular pathologies and interventions. A negative correlation between ΔAVAD and baseline AVAD was observed in this study. In other words, those with the lowest baseline AVAD had the greatest improvement in hydraulic force following AVR. Following removal of the increased afterload with AVR, reverse myocardial tissue remodelling takes place within the first year[9]. Prior research has demonstrated that post AVR there is a more pronounced regression in LVMi and diffuse myocardial fibrosis in those with the highest values of these parameters at baseline [29,30]. However, in these studies, higher LVMi at baseline was not shown to correlate with geometric reverse remodelling following AVR[29].

A more recent publication has hypothesized that a lack of geometric improvement seen in some individuals post AVR is attributable to the presence of myocardial fibrosis[31]. The findings of the present study indicate that the most marked improvement in AVAD post AVR occurs in those the poorest baseline AVAD, but the least severe myocardial tissue remodelling phenotype (lower LVMi, ECV), supporting this hypothesis. This suggests that early intervention in AS may be beneficial to allow improvement in hydraulic force before marked diffuse myocardial fibrosis and hypertrophy occur.

Baseline AVAD did not associate with baseline 6MWT, likely related the fact that all subjects had severe AS at baseline. However, post-operative AVAD was positively associated with post-operative functional capacity. Therefore, those with the best hydraulic force following AVR also had the best functional outcome.

### Limitations

Circular approximation was used to calculate the ASA, as opposed to measuring the true area, which will limit the accuracy of the ASA component used for the AVAD. Ideally ASA would be measured using the short axis acquisitions using the same machine learning model as VSA. However, the CMR protocol used to acquire short-axis images did not fully cover the left atria and therefore could not be used to assess the maximum ASA. AVAD was only measured during mid-diastole in this study. The hydraulic force of LV filling acts throughout the cardiac cycle and changes as the geometric relationship between the left atrium and left ventricles changes. Previous studies have measured the net hydraulic force throughout the cardiac cycle to demonstrate it in a small number of participants [18]. Mid-diastole was deemed to be the most useful single timepoint to measure the hydraulic force as it is when it has the greatest proportional impact on LV filling. This is in light of active relaxation and elastic restorative forces occurring in early diastole, and atrial contraction in late diastole.

## CONCLUSIONS

This is the first study assessing the hydraulic force of LV filling in a cohort of participants with severe aortic stenosis undergoing AVR. There was a net positive hydraulic force assisting left ventricular filling prior to and following valve replacement. Those with the most negative AVAD at baseline have the greatest improvement in hydraulic force following intervention. However, this improvement in hydraulic force is most marked in those yet to develop severe myocardial tissue remodelling, supporting the benefits of early intervention.

## ABBREVIATIONS

6MWT: 6-Minute Walk Test
AR: Aortic Regurgitation
AS: Aortic Stenosis
ASA: Atrial Short-axis Area
ASD: Atrial Septal Defect
AV: Atrioventricular
AVA: Aortic Valve Area
AVAi: Aortic Valve Area indexed to body surface area
AVAD: Atrioventricular Area Difference
AVADi: Atrioventricular Area Difference indexed to body surface area
AVR: Aortic Valve Replacement
CABG: Coronary Artery Bypass Graft
CMR: Cardiovascular Magnetic Resonance
ΔAVAD: change in Atrioventricular Area Difference
HFrEF: Heart Failure with reduced Ejection Fraction
hsTNT: high-sensitivity Troponin T
LA: Left Atrium
LGE: Late Gadolinium Enhancement
LV: Left Ventricle
LVEF: Left Ventricular Ejection Fraction
LVEDV: Left Ventricular End Diastolic Volume
LVEDVi: LV End Diastolic Volume indexed to body surface area
LVMi: Left Ventricular Mass indexed to body surface area
MR: Mitral Regurgitation
NT-pro BNP: N-terminal pro-brain natriuretic peptide
SAVR: Surgical Aortic Valve Replacement
TAVI: Transcatheter Aortic Valve Implantation
VSA: Ventricular Short-axis Area

## ACKNOWLEDGEMENTS

We gratefully acknowledge the contributions of the administrative and nursing staff, echocardiographers, radiographers, and biomedical scientists at the Barts Heart Centre, and University College London Hospitals.

## DATA AVAILABILTY

The data for this study is available from the corresponding author upon reasonable request.

